# Hepatitis C risk score as a tool to identify infected individuals: A demonstration study in Egypt

**DOI:** 10.1101/2023.07.10.23292449

**Authors:** Rayane El-Khoury, Hiam Chemaitelly, Ahmed S. Alaama, Joumana G. Hermez, Nico Nagelkerke, Laith J. Abu-Raddad

## Abstract

Hepatitis C virus (HCV) infection is a global health challenge. By the end of 2021, WHO estimated that less than quarter of global HCV infections were diagnosed. There is a need for a public health tool that can facilitate identification of infected persons and linking them to testing and treatment. We derived and validated a risk score to identify infected persons in Egypt and provided a demonstration of its utility. The 2008 and 2014 Egypt Demographic and Health Surveys were used to derive two risk scores using multivariable logistic regression. Both scores showed similar dependence on sex, age, and type of place of residence. Both risk scores demonstrated high and similar areas under the curve of 0.77 (95% CI: 0.76-0.78) and 0.78 (95% CI: 0.77-0.80), respectively. For the 2008 Risk Score, sensitivity was 73.7%, specificity was 68.5%, positive predictive value (PPV) was 27.8%, and negative predictive value (NPV) was 94.1%. For the 2014 Risk Score, sensitivity was 64.0%, specificity was 78.2%, PPV was 22.2%, and NPV was 95.7%. Each score was validated by applying it to a different survey database than the one used to derive it. Implementation of HCV risk scores is an effective strategy to identify carriers of HCV infection and to link them to testing and treatment at low cost to national programs.

## Introduction

Hepatitis C virus (HCV) infection is a global public health challenge^1, 2^ and a major cause of morbidity and mortality, resulting in liver cancer, fibrosis, and cirrhosis^3^. By end of 2021, the World Health Organization (WHO) estimated that 58 million people were infected with HCV, but only 15 million of them were diagnosed and only 9 million received treatment^4^. Direct-acting antivirals (DAA) offer highly effective treatment to cure this infection and to prevent progression toward severe forms of liver disease^5^, as well as an opportunity to reduce HCV transmission through treatment as prevention^6, 7^. Accordingly, the WHO has set a global target to eliminate HCV infection as a public health problem by 2030^2, 8^.

While DAAs are becoming accessible globally, it has been challenging to identify carriers of this infection so as to treat them, especially in the Middle East and North Africa (MENA), the global region most affected by HCV infection^9,10^. While mass testing and treatment programs may be relevant in high prevalence countries, other countries have relatively low HCV prevalence making such programs less cost-effective. There is a need for a public health tool that can assist in identifying potentially infected persons so as to link them to testing and treatment.

One such tool is the use of risk scores to identify potentially infected individuals. A risk score comprises a small set of simple questions that can be used to assess the likelihood that an individual has a specific health condition^11–14^, in this case, HCV infection. Such risk scores have proven influential as public health tools for a range of health conditions, such as diabetes^11–14^. Here, we demonstrate this public health tool for Egypt, to illustrate the utility of this concept.

## Methods

### Egypt Demographic and Health Surveys

The Egypt Demographic and Health Survey (EDHS) is a national survey that collected data pertaining to the health and demographics of a nationally representative sample of the resident population of Egypt, including HCV infection^15, 16^. The EDHS that included HCV biomarkers was conducted in 2008 and 2014 and used rigorous sampling methods^17^. Details on study design, data collection, and laboratory methods can be found in El-Zanaty et al.^15, 16^.

HCV antibody testing was done using a third generation enzyme-linked immunosorbent assay (ELISA), the Enzyme Immunoassay Adlatis EIAgen HCV Ab test (Adaltis Inc., Montreal, Canada)^15, 16^. All samples that were positive in the ELISA assay and 5% of the negative samples were then retested using a more specific assay, the chemiluminescent microplate immunoassay (CMIA ARCHITECT plus i1000SR, Abbott Diagnostic, USA)^15, 16^. If a sample was positive in both the ELISA and the CMIA testing, it was also tested for current active infection, using real-time, reverse-transcription polymerase chain reaction (RT-qPCR) testing to detect HCV ribonucleic acid (RNA)^15, 16^. Samples were further retested for internal and external quality assurance^15, 16^. Here we restrict our analyses to the HCV antibody results.

In the 2008 EDHS, 11,126 individuals 15-59 years of age were tested, of whom 1,571 were antibody positive^15^. The 2014 EDHS included children 1-14 years of age in addition to adults 15-59 years of age^16^. In this latter survey, 26,047 individuals were tested of whom 1,456 were antibody positive^16^.

Data from the EDHS 2008 and EDHS 2014 were downloaded with permission from Measure DHS^18^. For purposes of this study, the EDHS individual database was merged with the HCV biomarker database, based on established guidelines for managing DHS data^17^. All individuals with results for HCV antibody testing were included in the analysis.

### Risk score derivation

Associations of HCV antibody positivity (seropositivity) with a priori variables that are easy to evaluate in a primary care setting, and that can be included in a risk score, were investigated. These variables included sex (male versus female), age (5-year age strata), and type of place of residence (urban versus rural). Frequency distributions were generated to describe demographic and clinical profiles of tested individuals.

Chi-square tests and univariable logistic regression were implemented to investigate associations. Participants younger than 15 years of age were excluded as this age group was not included in the EDHS 2008 and has low HCV prevalence (Table 1)^6, 19–21^. Odds ratios (ORs), 95% confidence intervals (CIs), and p-values were reported. Covariates with p-values ≤0.1 in univariable regression analysis were considered possibly associated with HCV seropositivity. These were included in the multivariable analysis for estimation of adjusted odds ratios (AORs) and associated 95% CIs and p-values. Covariates with p-values ≤0.05 in the multivariable model were considered predictors of HCV seropositivity. Univariable and multivariable analyses were adjusted for sampling weights.

**Table 1.** Characteristics of individuals tested for HCV antibodies in the EDHS 2008 and 2014.

| Characteristics | EDHS 2008 |  |  | EDHS 2014 |  |  |
| --- | --- | --- | --- | --- | --- | --- |
|  | Total tested (%) | HCV antibody positive (proportion %) | p-value | Total tested (%) | HCV antibody positive (proportion %) | p-value |
| No | 11,126 | 1,571 |  | 26,047 | 1,456 |  |
| **Sex** |  |  |  |  |  |  |
| Female | 6,052 (54.4) | 711 (11.8) | <0.001 | 13,707 (52.6) | 660 (4.8) | <0.001 |
| Male | 5,074 (45.6) | 860 (17.0) |  | 12,340 (47.4) | 796 (6.4) |  |
| **Age group (years)** |  |  |  |  |  |  |
| 1-4 | - | - | - | 3,282 (12.6) | 10 (0.3) | <0.001 |
| 5-9 | - | - | - | 3,601 (13.8) | 10 (0.3) |  |
| 10-14 | - | - | - | 3,161 (12.1) | 23 (0.7) |  |
| 15-19 | 2,000 (18.0) | 82 (4.1) | <0.001 | 2,568 (9.9) | 30 (1.2) |  |
| 20-24 | 1,837 (16.5) | 91 (5.0) |  | 1,976 (7.6) | 54 (2.7) |  |
| 25-29 | 1,520 (13.7) | 92 (6.1) |  | 2,358 (9.0) | 88 (2.7) |  |
| 30-34 | 1,244 (11.2) | 133 (10.7) |  | 2,076 (8.0) | 114 (5.5) |  |
| 35-39 | 1,141 (10.3) | 147 (12.9) |  | 1,853 (7.1) | 130 (7.0) |  |
| 40-44 | 1,069 (9.6) | 238 (22.3) |  | 1,468 (5.6) | 146 (10.0) |  |
| 45-49 | 939 (8.4) | 275 (29.3) |  | 1,380 (5.3) | 208 (15.1) |  |
| 50-54 | 728 (6.5) | 272 (37.4) |  | 1,334 (5.1) | 337 (25.3) |  |
| 55-59 | 648 (5.8) | 241 (37.2) |  | 990 (3.8) | 306 (30.9) |  |
| **Type of place of residence** |  |  |  |  |  |  |
| Urban | 4,448 (40.0) | 442 (9.9) | <0.001 | 11,955 (45.9) | 546 (4.6) | <0.001 |
| Rural | 6,678 (60.0) | 1,129 (16.9) |  | 14,092 (54.1) | 910 (6.5) |  |

Abbreviations: EDHS, Egypt Demographic and Health Survey; HCV, Hepatitis C virus.

A risk score was constructed based on the β-coefficients obtained from the multivariable regression model. β-coefficients were multiplied by a factor of ten and then rounded to the nearest integer. The total risk score was calculated by adding the individual scores. To keep the score simple enough for use in primary care, we did not consider any interaction terms.

### Performance and validation of the risk score

A receiver operating characteristics (ROC) curve was plotted to investigate the performance of the risk score in predicting HCV seropositivity at different score cut-offs. A larger area under the curve (AUC), also called the c-index, indicates better performance of the risk score. The cut-off for the score was determined by maximizing the sum of the sensitivity and specificity. Sensitivity is the probability that the risk score will yield a positive diagnosis in a subject who is truly HCV antibody-positive. Specificity is the probability that the risk score will yield a negative diagnosis in a subject who is truly HCV antibody-negative.

Performance of the risk score was also investigated by estimating the positive predictive value (PPV) and the negative predictive value (NPV) of the risk score. PPV is the probability that a subject with a positive diagnosis per the risk score is truly HCV antibody-positive. NPV is the probability that a subject with a negative diagnosis per the risk score is truly HCV antibody-negative. The proportion of subjects who have scores greater than or equal to the cut-off of the risk score was estimated to determine the proportion of individuals that need to be biochemically tested for HCV antibodies.

To validate the performance of the derived EDHS 2008 risk score, the score was applied to the EDHS 2014 data to provide an independent validation using a different survey database than the one used to derive it. Here, the age variable was shifted to accommodate the 6-year aging between 2008 and 2014. Different performance diagnostics were then assessed. The same approach was also used to validate the EDHS 2014 risk score—it was applied to the EDHS 2008 database and performance diagnostics were assessed. Analyses were conducted in Stata version 16.1 (Stata Corporation, College Station, TX, USA).

## Results

Characteristics of individuals who were tested for HCV antibodies and the proportion of each population stratum that was HCV antibody-positive are shown in Table 1 for both of the EDHS surveys. Results of both surveys were consistent, taking into account the age shift in the national cohort with the passage of 6 years between the EDHS 2008 and EDHS 2014. HCV epidemiology in Egypt shows a strong age-cohort effect^6, 19–21^, thus the difference in HCV prevalence by age group in 2008 compared to 2014.

HCV seropositivity was strongly associated with sex, age, and place of residence in both national surveys (Table 2). Male sex and rural residence were associated with higher seropositivity. Seropositivity increased rapidly with age.

**Table 2.**
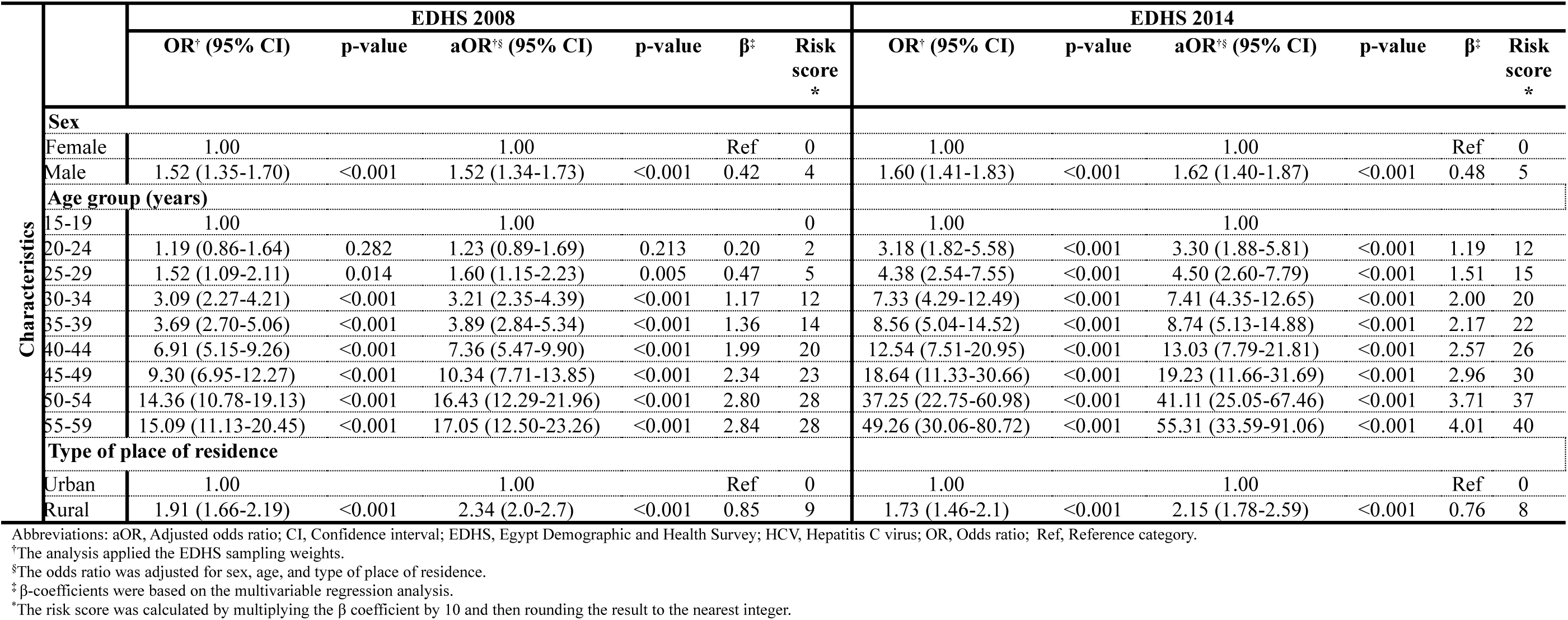
Results of univariable and multivariable regression analyses to derive the Egypt Hepatitis C Risk Score using data from EDHS 2008 and EDHS 2014.

The 2008 and 2014 Egypt Hepatitis C Risk Scores derived using the EDHS 2008 and EDHS 2014 data, respectively, are shown in Figure 1. The 2008 Risk Score had a range of 0-41. The 2014 Risk Score had a range of 0-53. Both showed similar dependence on sex, age, and type of place of residence. Both demonstrated high and similar AUCs (Figure 2). The AUC was 0.77 (95% CI: 0.75-0.78) for the 2008 Risk Score and 0.78 (95% CI: 0.77-0.80) for the 2014 Risk Score. The optimal combination of sensitivity and specificity was obtained at a score cut-off value of 22 for the 2008 Risk Score and at a cut-off of 34.5 for the 2014 Risk Score.

**Figure 1.**
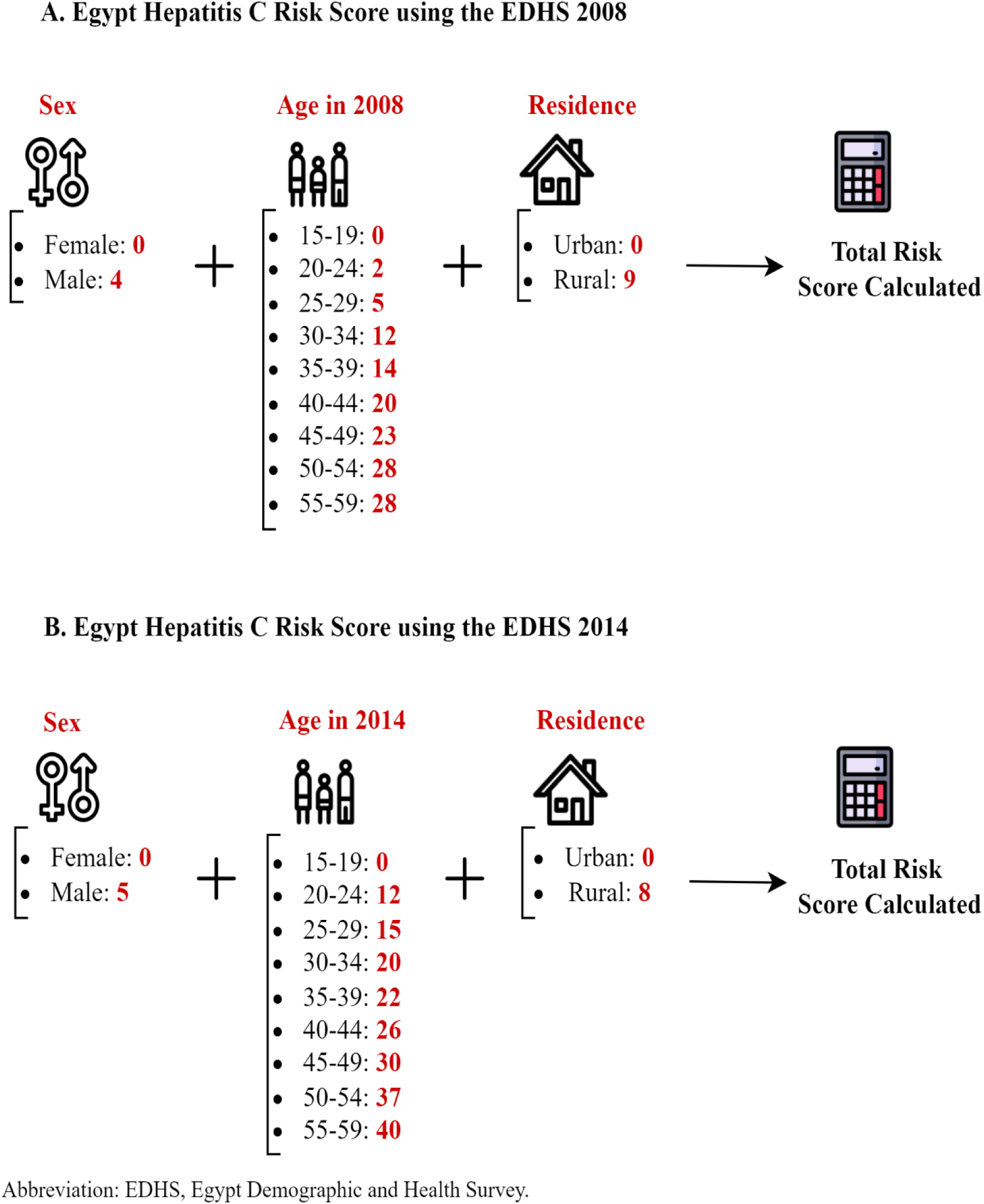
Mathematical formula of the derived Egypt Hepatitis C Risk Score.

**Figure 2.**
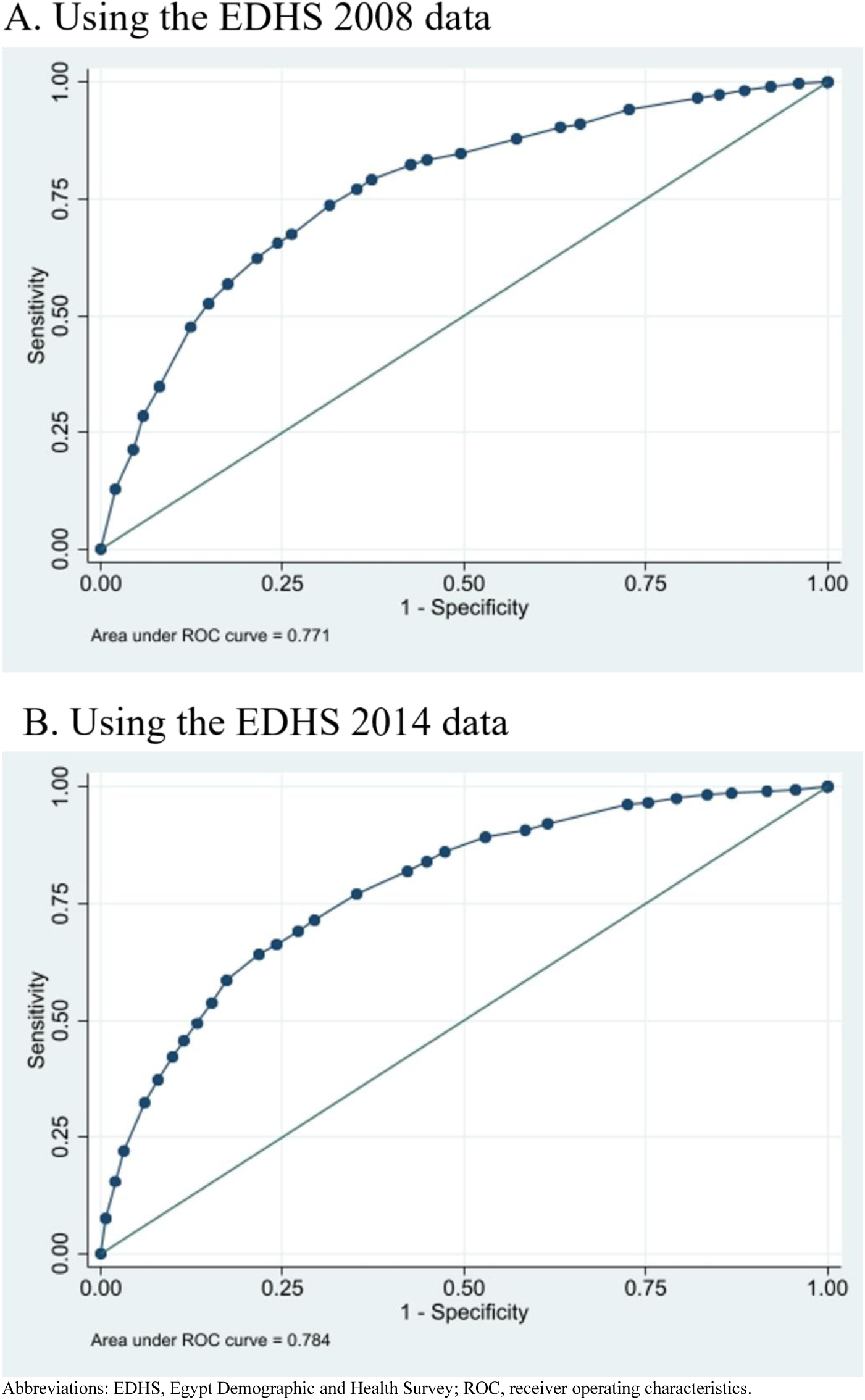
Diagnostic performance of the Egypt Hepatitis C Risk Score using the area under the (ROC) curve.

For the 2008 Risk Score, sensitivity was 73.7%, specificity was 68.5%, PPV was 27.8%, and NPV was 94.1% (Table 3). For the 2014 Risk Score, sensitivity was 64.1%, specificity was 78.2%, PPV was 22.2%, and NPV was 95.7%. The proportion of the population 15-59 years of age that needed to be biochemically tested for HCV antibodies was 37.2% using the 2008 Risk Score and 25.5% using the 2014 Risk Score. Of all HCV-infected persons in the EDHS samples, application of this score would have diagnosed (that is identified) 73.7% and 64.0% of all HCV antibody-positive persons in samples of the EDHS 2008 and 2014, respectively.

**Table 3.**
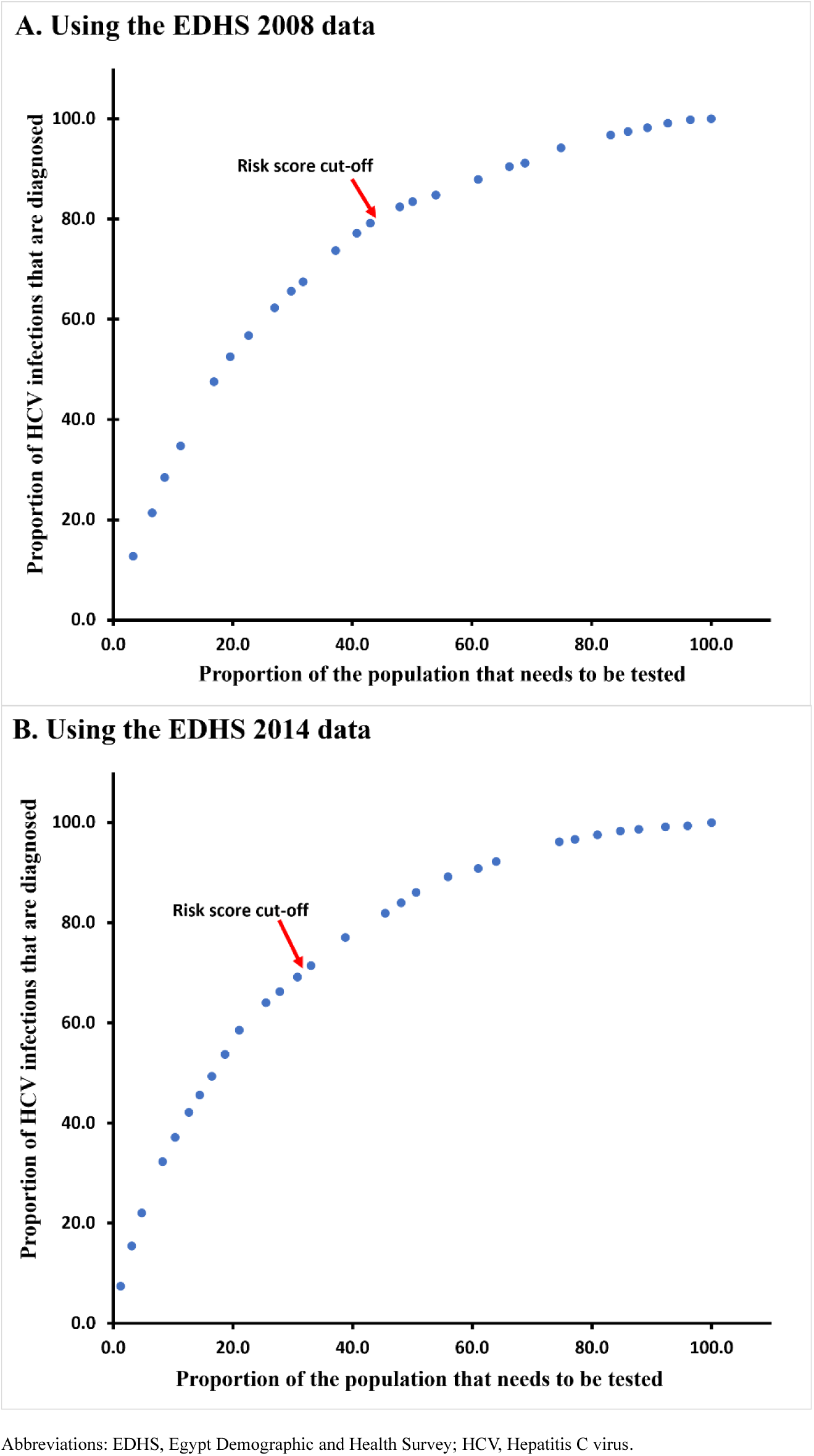
Performance of the Egypt Hepatitis C Risk Score.

When the 2008 Risk Score was applied to the EDHS 2014 data, the AUC was 0.75, the sensitivity was 66.1%, and the specificity was 72.3% (Table 3). These performance indicators were similar to the original performance indicators generated using the EDHS 2008 data. Therefore, this application validates this risk score. A similar outcome was found when the 2014 Risk Score was applied to the EDHS 2008 data, also providing a validation of the 2014 risk score.

Figure 3 displays the proportion of HCV infections in the population that are diagnosed as a function of the proportion of the population that needs to be tested to identify these infections, using each of EDHS 2008 and EDHS 2014 data. The figure shows the effect of prioritization of testing for those with higher to lower risk score. This provides a demonstration of the utility of using the risk score: a large proportion of HCV infections can be diagnosed by testing only a small proportion of the population. It is most efficient programmatically to start testing individuals with the highest risk score and progressively moving on to those with lower and lower risk scores. As testing is expanded to those with low risk scores, the yield in identifying more HCV infections is very limited.

**Figure 3.**
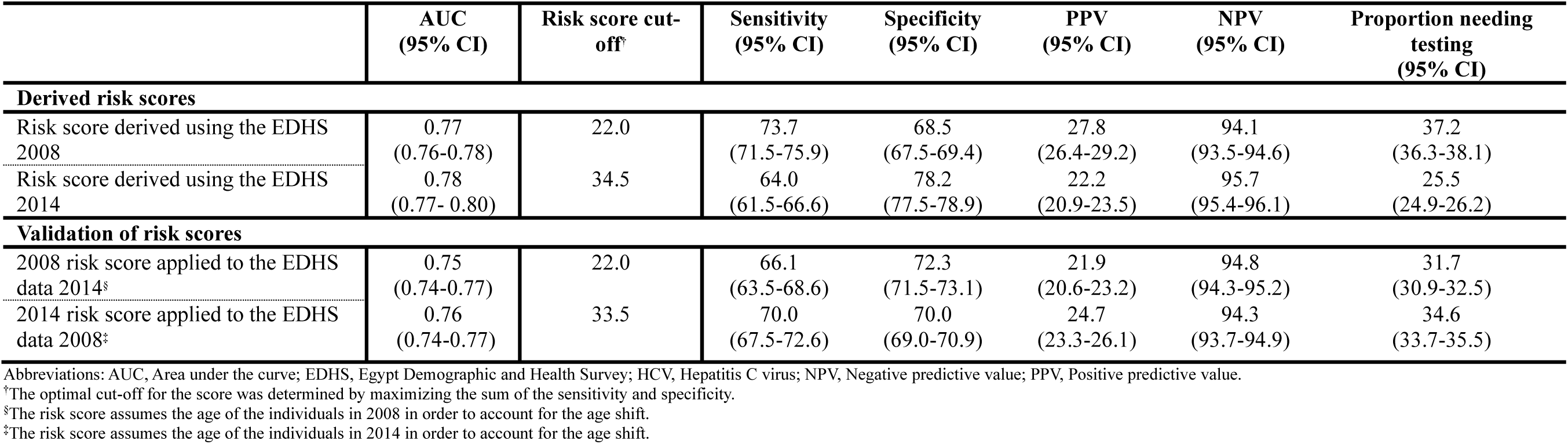
Proportion of HCV infections in the population that are diagnosed as a function of the proportion of the population that needs to be tested to identify these infections. The figure shows the effect of prioritization of testing for those with higher to lower risk score.

## Discussion

We demonstrated that a risk score that consists of few simple questions that are easy to evaluate in a primary care setting or implemented through a website or an app that helps persons identify their risk of being HCV-infected, provides an effective and non-invasive public health tool to identify carriers of HCV infection and to link them to testing and treatment. Biochemical testing methods to identify HCV infected persons are invasive and time-consuming and require human and financial resources, as well as complex logistics, making them less scalable, particularly in resource-limited settings. In contrast, initial screening using a risk score can be easily administered or self-administered, is non-invasive, and requires minimal resources and logistics. Therefore, HCV risk scores can be an indispensable strategy for the global response to attain the target of HCV elimination as a public health problem by 2030^2, 8^. HCV risk scores may also prove useful in disseminating awareness about this infection in the wider population.

Remarkably, the risk score, which consists of three simple questions, provided considerable diagnostic accuracy, as is evident in the values of various diagnostic metrics, including AUC, sensitivity, specificity, PPV, and NPV. It is striking that such a simple risk score managed to achieve diagnostic accuracy comparable to some of the biochemical assays used in healthcare practice for a range of diseases. The score was also able to identify 73.7% and 64.0% of all HCV infections in the EDHS 2008 and EDHS 2014 samples, respectively. This further demonstrates the value of this approach to identify as many HCV-infected persons as early as possible, and treating them before progression to serious clinical disease.

This approach was demonstrated for Egypt, considering the availability of two EDHS surveys to derive and validate the score. The two scores showed similar structures and similar diagnostic performances. Each score was validated by applying it to a database other than the one used to derive it. The latter application yielded a diagnostic performance that was comparable to the original diagnostic performance against the database used to originate it. This highlights how a single national survey for HCV infection may be sufficient to develop an effective risk score for this infection, and that can become an integral component of the national response to eliminate HCV.

While this study focused on demonstrating the utility of this concept as a public health tool, actual application of this approach to different countries can be enhanced for even higher diagnostic accuracy. One extension could be adding more variables to the score in a manner tailored to the local epidemiology of each country. For instance, province or city of birth and/or current residence, prior exposure to an HCV mode of transmission^22^, or history of HCV infection in the family, could be added, among others. Given that the risk of exposure to HCV infection varies immensely by at-risk subpopulation type and shows a distinctive hierarchy^23^, an additional component to the score could be to integrate the at-risk subpopulation type as a variable^23, 24^, thereby further enhancing the diagnostic accuracy of the score. Testing strategies, therefore, could be highly efficient in identifying HCV infected persons at a modest cost.

This study has limitations. For ease of use in primary care and more broadly by the public, a risk score has to be simple. Accordingly, it cannot fully represent the complex epidemiology of HCV infection, such as interactions among risk factors. This risk score was derived for Egypt, which may not benefit from this risk score, given that this country has opted for mass testing of its entire population^25^. Derivation of a risk score requires at least one round of a population-based survey, ideally at the national level, but many countries may not have such survey data to be able to derive the risk score. The risk score was derived for a high-burden country, and utility of this approach still needs to be demonstrated for countries with low HCV prevalence. Nonetheless, this approach may prove to have higher utility in countries with low HCV prevalence than in countries with high HCV prevalence, as HCV epidemiology shows a clearer hierarchy in infection exposure risk in countries with concentrated HCV epidemics compared to those with generalized HCV epidemics^23^.

## Conclusions

An HCV risk score can be derived using only one round of a population-based survey and offers an effective, simple, non-invasive strategy to identify carriers of HCV infection and to link them to testing and treatment, at low cost. This public health tool can be implemented and used for prioritizing sub-populations for interventions with minimal logistical complexity and cost, especially in resource-limited countries.

## Author’s contributions

REK conducted the data analyses with HC and NN. REK, NN, and LJA co-wrote the first draft of the article. LJA conceived and led the design of the study, analyses, and drafting the article. All authors contributed to drafting and revising the manuscript. All authors have read and approved the final manuscript.

## Availability of Data and Materials

All data analyzed in this study can be accessed through application to the DHS Program at https://dhsprogram.com/ or by contacting.

